# Hospital *Enterococcus faecium* demonstrates distinct environmental and patient reservoirs: a genomic point prevalence survey

**DOI:** 10.1101/2024.10.09.24315211

**Authors:** Nenad Macesic, Hugh Cottingham, Jessica A. Wisniewski, Luke V. Blakeway, Ravali Theegala, Katherine Pragastis, Andrew Stewardson, Pauline Bass, Megan Gritt, Stephanie Spilsbury, Denise Del Rosario-Kelly, Amanda Dennison, Denis W. Spelman, Adam W.J. Jenney, Anton Y. Peleg

## Abstract

**Objectives:** *Enterococcus faecium* is a common cause of hospital-acquired infections and is associated with vancomycin resistance. The hospital environment is often a reservoir for *E. faecium* due to its ability to survive on surfaces. In 2019 we noted increasing prevalence of *vanA E. faecium* causing bloodstream infections. We aimed to assess the hospital environment as a reservoir of *vanA E. faecium*.

**Methods:** We conducted a point prevalence survey of the environment at the Alfred Hospital, Melbourne, Australia. We sampled all wards (n=12), 10 surface types and 357 surfaces. Resulting *E. faecium* isolates underwent short-read whole genome sequencing and were compared to invasive infection (bloodstream) isolates. The *van* operon, multi-locus sequence type (MLST) and core genome MLST (cgMLST) were determined.

**Results:** Twenty percent (73/357) of surfaces isolated *E. faecium* with a median of 20% positivity per ward (IQR 11.6-30.6%). *vanA* and *vanB* were detected in 12/73 (16%) and 58/73 (79%) environmental genomes, respectively. In contrast, a higher proportion of bloodstream isolates carried *vanA* (32/72 [44%], *P*<0.001). Environmental genomes belonged to 10 MLSTs and 10 cgMLSTs, with ST796/ST78 accounting for 54/73 (74%) genomes. Clinical STs and cgMLSTs overlapped with environmental but were more diverse. We noted 13 putative genomic transmission clusters. One cluster contained 25/44 (57%) *vanA* genomes, with the majority being clinical (22/26 genomes, 85%). Environmental-clinical links were observed in 292/895 (33%) putative genomic transmission links. In completed assemblies, the *vanA* and *vanB* operons were located on pRE25-like plasmids and the chromosome, respectively.

**Conclusion:** We noted a 20% prevalence of *E. faecium* environmental colonisation but the *vanA* operon was detected in only 12/73 genomes. We identified distinct environmental and patient reservoirs, with most environmental genomes carrying the *vanB* operon and having distinct STs/cgMLST clusters. Environment-clinical reservoir spillover was detected but accounted for a minority of putative transmission links.

## Introduction

*Enterococcus faecium* is a common cause of hospital-acquired infections and is frequently associated with vancomycin resistance [1], resulting in high mortality, longer hospital stays and higher healthcare costs [2]. The hospital environment is a reservoir for *E. faecium* due to its ability to survive on surfaces for prolonged periods [1]. Prior studies have analysed patient and environmental reservoirs of *E. faecium* but have typically been limited to selected hospital wards in settings where either *vanA* or *vanB* vancomycin-resistant *E. faecium* (VREfm) predominate [3-5]. Australia has a specific VREfm epidemiology with more recent emergence of *vanA* VREfm on a background of *vanB* VREfm endemicity, with both continuing to circulate [6]. In 2019, we noted increasing prevalence of *vanA* VREfm bloodstream infections. We aimed to assess the hospital environment as a reservoir of *vanA* VREfm by conducting a point prevalence study and comparing environmental and contemporary clinical isolates using genomic analyses.

## Methods

The study was approved by the Alfred Hospital Ethics Committee. The Alfred Hospital (Melbourne, Australia) is a 600-bed quaternary hospital with state referral services for burns and trauma, as well as stem cell and solid organ (heart/lung/kidney) transplant services. We conducted a point prevalence survey of the hospital environment. We sampled surfaces using FLOQswabs (Copan) over two months, which included all wards and the intensive care unit (ICU) (n=12), with 10 surface types sampled per ward/ICU. Samples were cultured on CHROMagar VRE media (Biomerieux) and species were identified using matrix-assisted laser desorption-ionisation time-of-flight (MALDI-TOF) (Bruker Daltonics). To assess for molecular epidemiological links between environmental and clinical isolates, we selected all *E. faecium* blood culture isolates from our established biobank from 6 months prior to 6 months post the environmental sampling (Supp. Figure 1 and Supp. Table 1). Clinical data were extracted from the electronic medical record.

All *E. faecium* isolates underwent short-read (Illumina) whole genome sequencing, as described previously [7]. We detected *van* operon presence and assigned multi-locus sequence type (MLST) and core genome MLST (cgMLST) [8]. For each cgMLST cluster, we assembled a long-read reference genome and calculated within-cluster pairwise single nucleotide variant (SNV) distances. Please see Supplementary Methods for full details.

## Results

### Environmental point prevalence survey

We swabbed 357 surfaces (median 31.5 per ward [range 10-60]). A total of 73/357 (20%) surfaces isolated *E. faecium*, with median 20% positive per ward (IQR 11.6-30.6%) (Fig. 1A), indicating that *E. faecium* was endemic in the hospital environment. Rates of *E. faecium* positivity varied widely, with the highest rate in the Burns ward (10/10 swabs positive) and the lowest rate on Surgical Ward 3 (0/34 positive). Patient chairs had the highest rate of *E. faecium* positivity (13/25 [52%]) (Fig. 1B).

**Figure 1.**
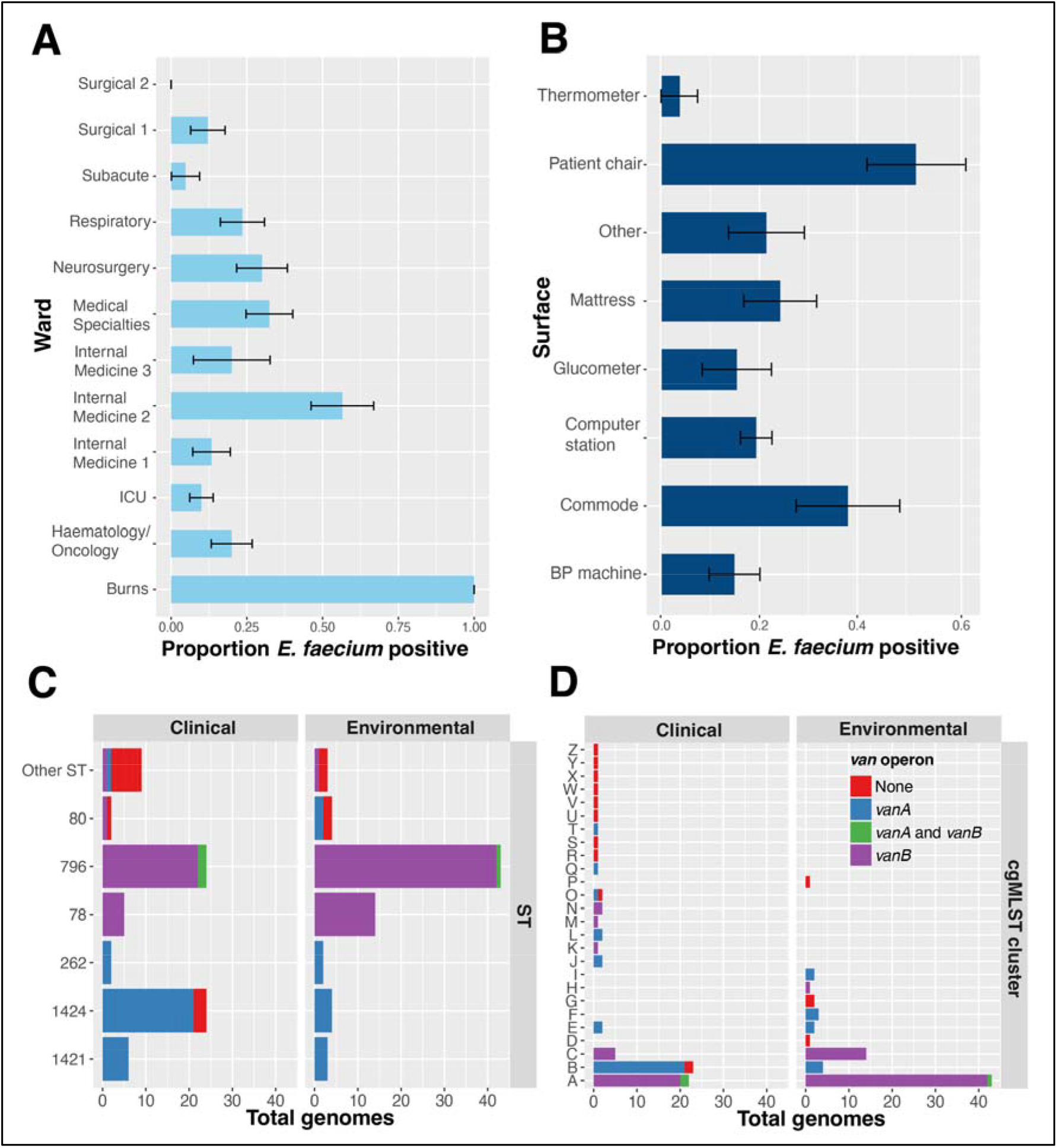
Summary of *Enterococcus faecium* environmental screening and contemporary clinical isolates. **A)** Proportion of environmental screening swabs positive for *E. faecium* by ward and **B)** by surface type. **C)** Summary of *E. faecium van* operon presence by multi-locus sequence types (MLST) and **D)** core genome MLST (cgMLST) in clinical and environmental isolates.

### Environmental and clinical genome characteristics

All 72 *E. faecium* blood culture isolates that occurred in the 6 months pre- and post-environmental sampling were sequenced and compared to the 73 positive environmental swabs (Fig. 1C, Supp Fig. 1). *vanA* and *vanB* were detected in 12/73 (16%) and 58/73 (79%) environmental genomes, respectively (Fig. 1C). In contrast, 32/72 (44%) clinical genomes carried *vanA* (*P*<0.001) and 31/72 (43%) carried *vanB*. Environmental genomes belonged to 10 MLSTs and 10 cgMLSTs, with ST796/ST78 accounting for 54/73 (74%) genomes (Fig. 1C, 1D). Clinical STs/cgMLSTs overlapped with environmental genomes but were more diverse. Specifically, ST1421/1424 carried *vanA* and were more frequent in clinical genomes (30/72 v 7/73 genomes, *P*<0.001). A single cgMLST cluster (B) contained the majority of *vanA* genomes (25/44 genomes), while *vanB* genomes were noted across two cgMLST clusters (A and C, 84/89 genomes) (Fig. 1D). STs/cgMLSTs carried either *vanA* or *vanB* operons, except for 3 genomes in ST796/cgMLST cluster A that had concurrent *vanA* and *vanB* carriage (Fig. 1C/1D).

### Geography of E. faecium on hospital wards

We mapped *E. faecium* ST/cgMLST and presence of *van* operon across individual wards (Supp. Fig. 2). High-risk wards (ICU, Haematology/Oncology) had the majority of clinical genomes with a resultant high diversity of STs/cgMLSTs (7 STs/10cgMLSTs for Haematology/Oncology and 7 STs/7 cgMLSTs for ICU, respectively). The Haematology/Oncology ward had the highest proportion of *vanA* genomes (12/25, 48%), with 7/12 of these being ST1424/cgMLST cluster B genomes. Other wards had uniform ST/cgMLST composition with no differences between wards (Supp. Fig. 2).

### E. faecium *transmission networks*

Using a 6 SNV cutoff [4], there were 895 putative genomic transmission links resulting in 13 clusters (Fig. 2). While 292/895 (33%) links were environmental-clinical, of these only 88/292 (30%) were between *vanA* genomes. Most clusters had a predominance of either environmental-environmental links or clinical-clinical links (Supp. Table 2). A single cluster contained 25/44 (57%) *vanA* genomes, with the majority being clinical (22/26, 85%). Environmental genomes in that cluster came from three wards, while clinical genomes came from 11 wards. In contrast, *vanB* genomes belonged to three major (>5 genomes) clusters, containing predominantly environmental genomes (42/56, 75%).

**Figure 2.**
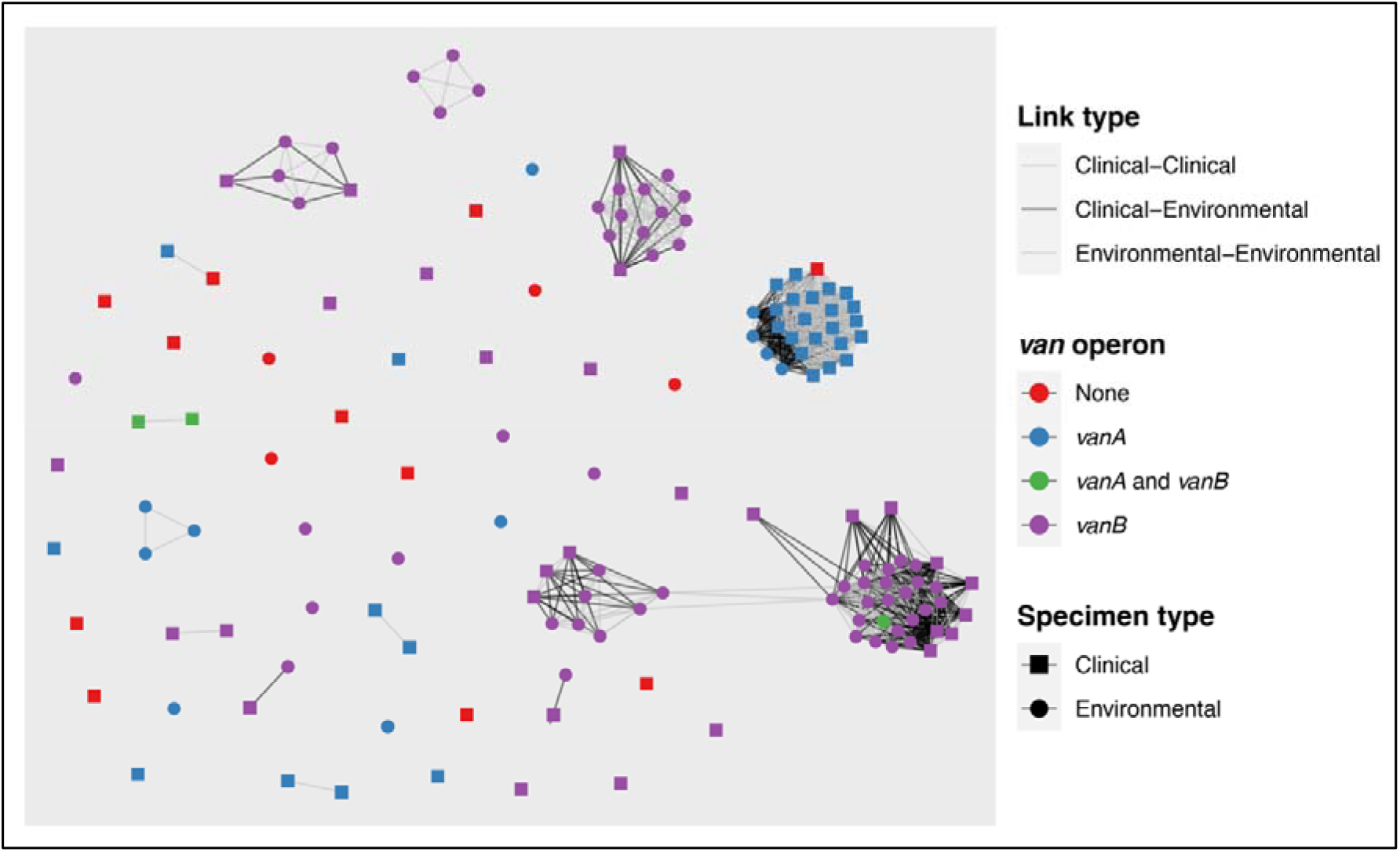
Network analysis of *Enterococcus faecium* putative genomic transmission links. Clinical and environmental *E. faecium* genomes are shown as nodes and putative genomic transmission events (defined as pairwise single nucleotide variant distance ≤6) as edges. Edges resulting from clinical-environmental links are shown in black, and clinical-clinical or environmental-environmental in grey. A single major *vanA* cluster is noted with majority of clinical genomes. In contrast, three major *vanB* clusters are noted with environmental genomes predominating.

### van *operon analysis*

In completed assemblies (n=26), the *<u>vanA</u>* and *vanB* operons were located on pRE25-like plasmids and the chromosome, respectively. The pRE25-like plasmids were heterogeneous and did not indicate spread of a single plasmid across multiple cgMLST clusters (Supp. Figure 3).

## Discussion

We noted a 20% prevalence of *E. faecium* environmental colonisation across a diverse range of hospital surfaces, but only 12/73 (17%) environmental genomes carried *vanA*. This indicated that while *E. faecium* was endemic in the hospital environment, it likely did not make a major contribution to the increased *vanA* VREfm prevalence in bloodstream infection isolates. Indeed, we noted distinct environmental and patient reservoirs: an environment dominated by *vanB* ST796/78 genomes, and a clinical reservoir comprising *vanA* ST1421/1424 genomes. There was evidence of spill-over with putative clinical-environmental links, but these were a minority (33%) and *vanA* putative genomic links only contributed 88/895 (10%) of these in turn.

Our findings stand in contrast to prior work showing significant links between VREfm in the environment and clinical colonisation and infection [3-5]. This difference may be due to these studies describing non-outbreak settings and focusing on limited wards (ICU and Haematology/Oncology). Potential contributors to our findings include our institution’s differing infection prevention approaches to *vanA* and *vanB* VREfm-colonised patients [9]: *vanA* patients are routinely placed in contact precautions, while for *vanB* patients this occurs only on high-risk wards and if they have diarrhoea or non-contained wounds. *E. faecium* ST796, a key *vanB* lineage in our study, has been shown to develop biocide tolerance that may provide it with a fitness advantage in hospital environments [10]. Of note, the *vanA* clinical reservoir that prompted our investigation subsequently resolved during the COVID-19 pandemic (Supp. Fig. 1), perhaps because of these factors.

Although the environment plays a major role in healthcare-associated VREfm spread, our study shows that distinct clinical and environmental reservoirs of VREfm may exist, particularly in outbreak settings where new clones are emerging and have not yet colonised the environment. This provides support for use of molecular techniques to focus infection prevention strategies according to the reservoir. While cleaning strategies aiming to reduce the environmental burden form an essential part of any multifaceted approach to VREfm control [11], the clinical reservoir may require more active attention through measures such as introduction of active surveillance, redoubled hand hygiene efforts and more stringent application of contact precautions in order to prevent both between-patient spread and future environmental colonisation.

## Data Availability

All data produced in the present study are available upon reasonable request to the authors

## Conference Presentation

Portions of this work were presented at European Society of Clinical Microbiology and Infectious Diseases (ESCMID) Global 2024, held in Barcelona, Spain.

