## Supplement for "Hospital *Enterococcus faecium* demonstrates distinct environmental and patient reservoirs: a genomic point prevalence survey"

**Supplementary Methods**

*Culture, DNA extraction and sequencing*

All bacterial isolates were grown on Brain Heart Infusion agar (Becton-Dickinson) at 37°C for 16 hours. Genomic DNA was extracted from bacterial plate culture using the GenFind V3 Reagent Kit (Beckman Coulter) as per manufacturer's instructions. Libraries for short read sequencing were prepared using the Nextera Flex DNA Library Prep Kit (Illumina), and 150 bp paired-end sequencing was performed on the NovaSeq 6000 system (Illumina). Libraries for long-read sequencing were prepared using the Ligation Sequencing Kit with Native Barcoding Expansion (Oxford Nanopore Technologies) and sequenced on the GridION instrument with an R10.4.1 flow cell (Oxford Nanopore Technologies) for 48 hours. Basecalling was performed with Guppy v.4.0.14 using the 'super' basecalling model.

*De novo assembly and annotation*

We constructed *de novo* assemblies of all isolates with only short-read data using Unicycler [1]. Assembly quality was checked using Quast v5.2.0 [2] and species identification was performed using GTDB-Tk v1.0.2 [3]. Genomes were annotated using Prokka v1.14.6 [4]. We then performed resistance gene and plasmid replicon detection with Abricate v.1.0.0 [5], using the NCBI Antibiotic Resistance and PlasmidFinder databases, respectively. We determined *in silico* multi-locus sequence type (ST) using 'mlst' v.2.19.0 [6].

*Core genome multi-locus sequence typing (cgMLST) and related analyses*

We defined cgMLST of each isolate using the approach of Higgs et al. [7]. In brief, we assigned cgMLST alleles using the public *Enterococcus faecium* cgMLST scheme [8] (core set of 1,423 genes) and chewBBACA (v2.0.16) [9]. We then determined the number of allelic differences between each genome and assigned cgMLST clusters using single linkage clustering and a pairwise allelic difference threshold of ≤25.

For each cgMLST cluster with ≥2 genomes, we then selected the earliest available isolate as a reference and generated long-read sequencing data as described above. We then constructed *de* *novo* assemblies incorporating short- and long-read data for these isolates. We used a long-read-first assembly approach using a bespoke pipeline (https://github.com/HughCottingham/clinopore-nf) that incorporates Flye v2.9.2 with subsequent polishing with Medaka v1.8.0, Polypolish v0.5.0 and Polca v3.4.1 [10-14].

Using these completed assemblies as references, we generated a core chromosomal single nucleotide variant (SNV) alignment using Snippy v.4.6.0 [15] for each cgMLST. We then calculated pairwise SNV distances for all genomes in that cgMLST. Genomes with ≤6 SNV differences were considered putative genomic transmission events [16]. We then used the R package 'ggraph' v2.0.5 to visualize putative transmission networks with genomes as nodes and putative genomic transmission events as edges.

*Plasmid analyses*

Using Abricate, we identified *van* operon-harboring contigs that were putative plasmids in our reference completed assemblies. We used progressiveMauve v2.4.0.r4736 to align all plasmids and assess for structural re-arrangements [17]. Geneious Prime 2023.2.1 (https://www.geneious.com) was used to visualize and assess for structural re-arrangements, with subsequent manual annotation in Adobe Illustrator v2020.24.3.

*Data availability*

Illumina/Nanopore read data were deposited in the NCBI Sequence Read Archive under project accession PRJNA1077967. Completed genome assemblies were deposited in GenBank; accessions are listed in Supp. Table 1.

*Statistical analysis*

Categorical variables were compared using $\chi 2$ or Fisher's exact tests and continuous variables were compared using Student's *t*-test or Mann-Whitney-Wilcoxon, as appropriate. Statistical analyses were performed in R (v4.3.0).

**Supp. Fig. 1 – *Enterococcus faecium* clinical bloodstream isolate collection**

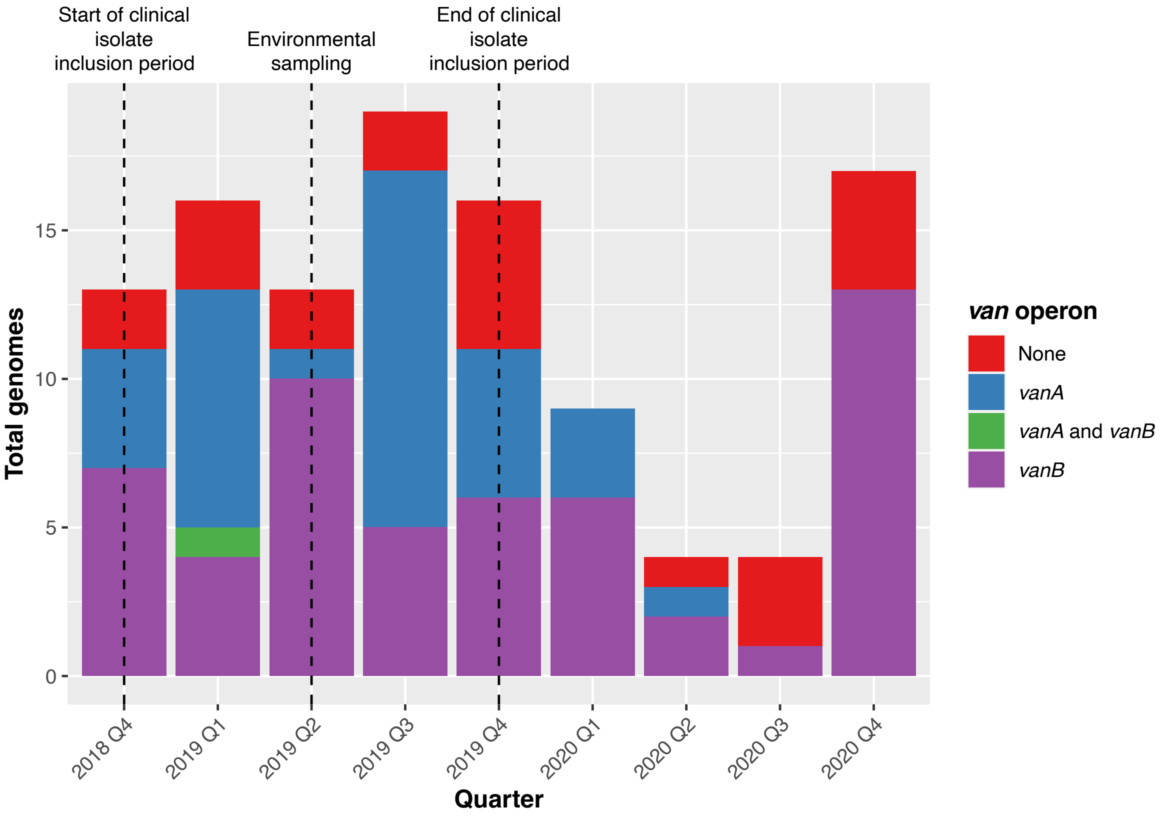


Summary of *E. faecium* bloodstream isolate collection by year and quarter, with presence of *van* operon indicated. Dotted lines show bloodstream isolate inclusion period and time of environmental sampling.

**Supp. Fig. 2 – Summary of *Enterococcus faecium* environmental and clinical genomes by ward**

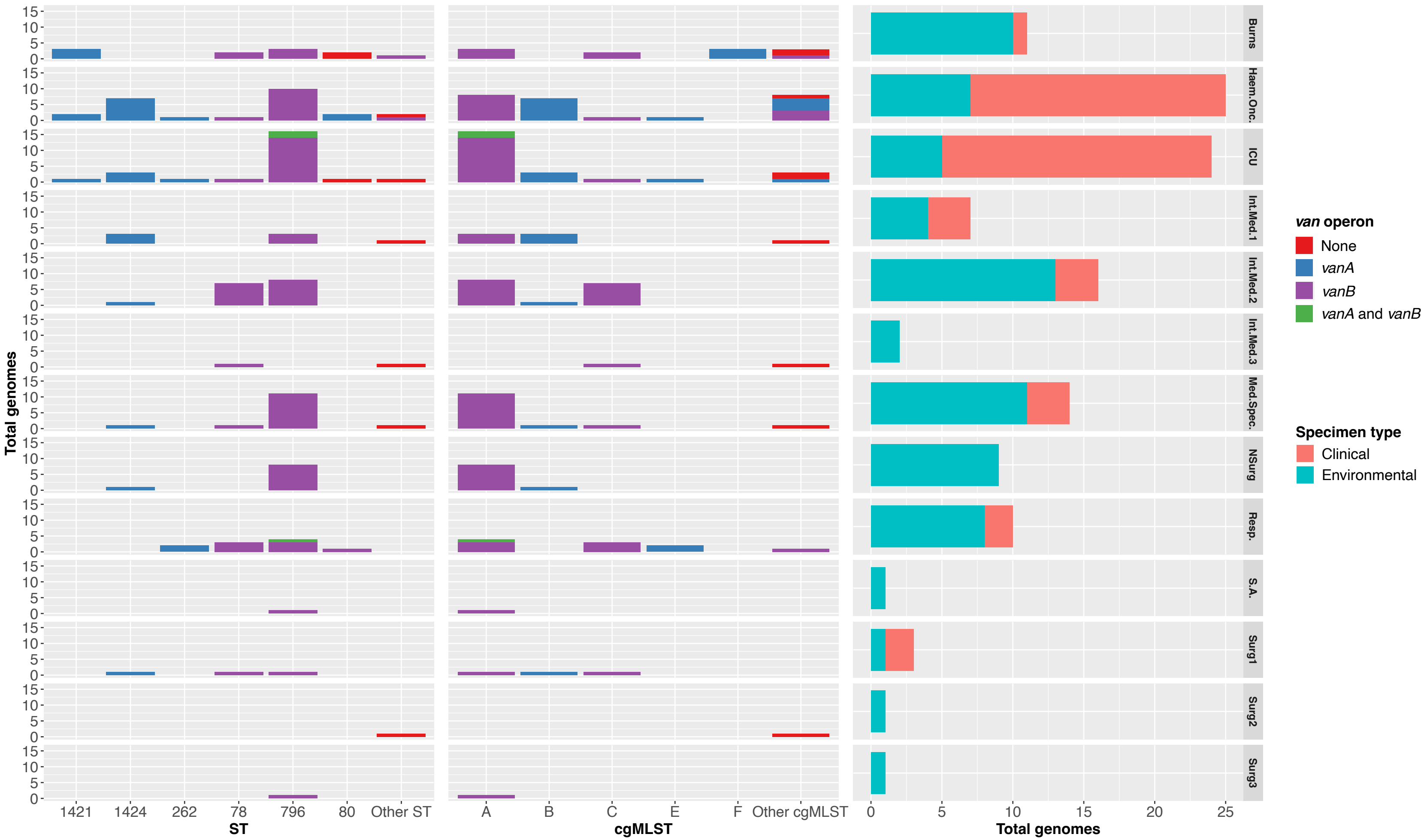


For each ward, environmental and clinical *E. faecium* genomes are shown by multi-locus sequence type (MLST) and core genome MLST (cgMLST). Abbreviations: Haem.Onc. – Haematology/Oncology; ICU – Intensive Care Unit; Int.Med. – Internal Medicine; Med.Spec. – Medical Specialties; NSurg – Neurosurgery; Resp. – Respiratory; S.A. – Subacute; Surg. – Surgery.

**Supp. Figure 3 – Plasmids with *van* operons from *Enterococcus faecium* completed assemblies**

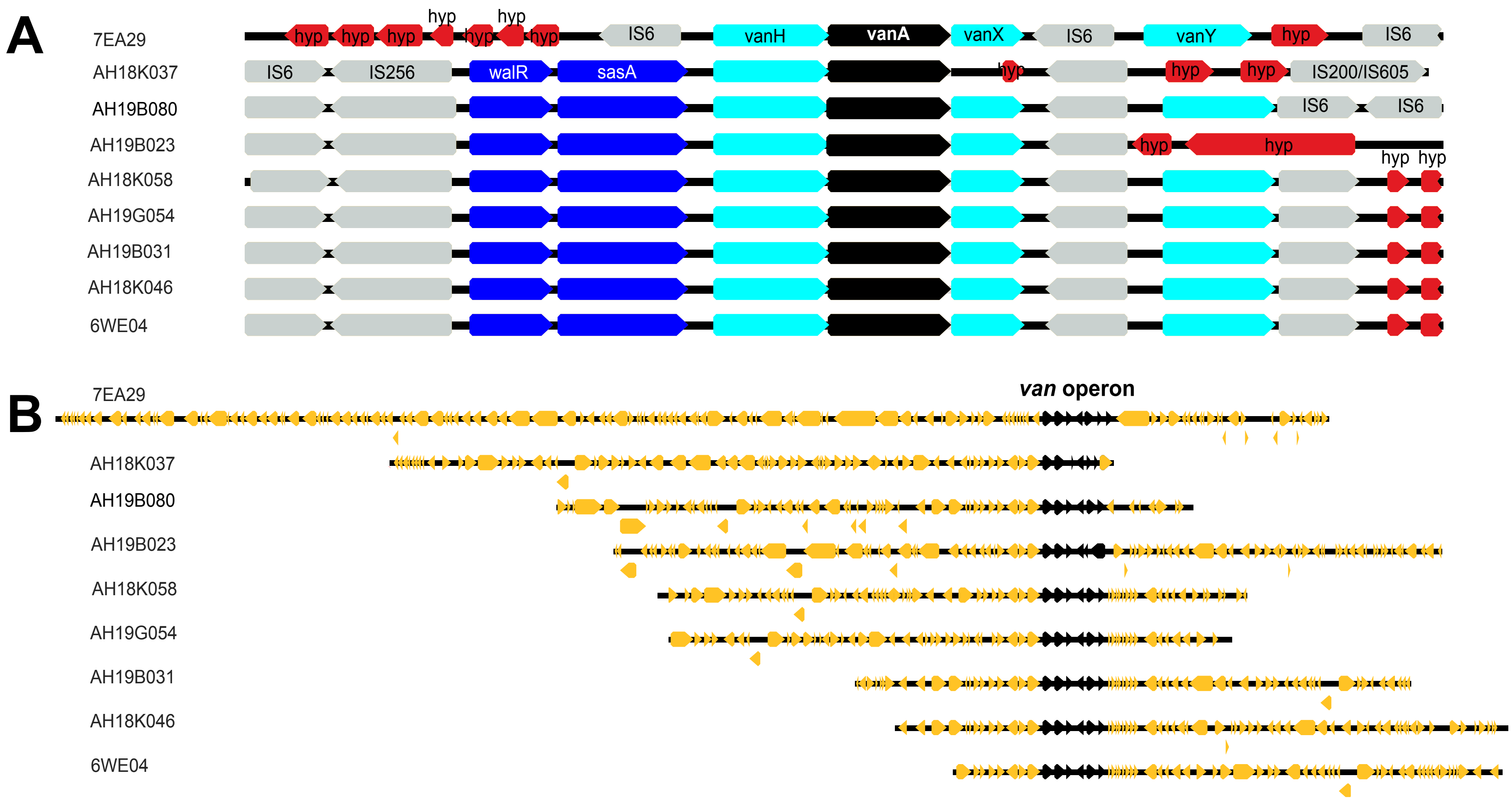


A) Detailed view of *van* operons within Tn1546 transposons from plasmids in completed *E. faecium* assemblies. Isolate numbers are shown in column on left. B) *van* operons (in black) shown in context of entire plasmids (yellow) from completed *E. faecium* assemblies. The plasmids carried additional resistance determinants including *aphA* (aminoglycosides), *erm* (macrolides), *tet(L)* and *tet(M)* (tetracyclines) and *bcrABR* (bacitracin).

**Supp. Table 1 - Summary of included clinical and environmental Enterococcus faecium genomes**

| Isolate | Collection Date | Specimen Type | *van* operon presence | ST | cgMLST cluster | SRA (Illumina) accession no. | SRA (Nanopore) accession no. | GenBank accession no. |
|---|---|---|---|---|---|---|---|---|
| 2EA01 | 12/6/2019 | Environmental | VanB | 796 | A | SRR28039592 | | JBAKTT000000000 |
| 2EA04 | 12/6/2019 | Environmental | VanA | 1424 | B | SRR28039591 | | JBAKTS000000000 |
| 2EA05 | 12/6/2019 | Environmental | VanB | 796 | A | SRR28039575 | | JBAKTR000000000 |
| 2EA09 | 12/6/2019 | Environmental | VanB | 796 | A | SRR28039564 | | JBAKTQ000000000 |
| 2EA15 | 12/6/2019 | Environmental | VanB | 796 | A | SRR28039451 | | JBAKTP000000000 |
| 2EA17 | 12/6/2019 | Environmental | VanB | 796 | A | SRR28039440 | | JBAKTO000000000 |
| 2EA21 | 12/6/2019 | Environmental | VanB | 796 | A | SRR28039429 | | JBAKTN000000000 |
| 2EA24 | 12/6/2019 | Environmental | VanB | 796 | A | SRR28039520 | | JBAKTM000000000 |
| 2EA32 | 12/6/2019 | Environmental | VanB | 796 | A | SRR28039509 | | JBAKTL000000000 |
| 2WE08 | 6/5/2019 | Environmental | VanB | 78 | C | SRR28039498 | | JBAKTK000000000 |
| 2WE22B | 6/5/2019 | Environmental | None | 17 | D | SRR28039590 | SRR28039475 | JBAKTJ000000000 |
| 2WE35 | 6/5/2019 | Environmental | VanB | 796 | A | SRR28039546 | | JBBDHB000000000 |
| 4EA04 | 19/6/2019 | Environmental | VanA | 1424 | B | SRR28039535 | | JBAKTI000000000 |
| 4EA05 | 19/6/2019 | Environmental | VanB | 796 | A | SRR28039524 | | JBAKTH000000000 |
| 4EA30 | 19/6/2019 | Environmental | VanB | 796 | A | SRR28039481 | | JBAKTG000000000 |
| 4EA31 | 19/6/2019 | Environmental | VanA | 1424 | B | SRR28039470 | | JBAKTF000000000 |
| 4WA01 | 6/5/2019 | Environmental | VanB | 796 | A | SRR28039459 | | JBAKTE000000000 |
| 4WA02 | 6/5/2019 | Environmental | VanB | 796 | A | SRR28039579 | | JBAKTD000000000 |
| 4WA03 | 6/5/2019 | Environmental | VanB | 78 | C | SRR28039577 | | JBAKTC000000000 |
| 4WA06 | 6/5/2019 | Environmental | VanB | 796 | A | SRR28039576 | | JBAKTB000000000 |
| 4WA07 | 6/5/2019 | Environmental | VanB | 796 | A | SRR28039574 | | JBAKTA000000000 |
| 4WA08 | 6/5/2019 | Environmental | VanB | 796 | A | SRR28039573 | | JBAKSZ000000000 |
| 4WA09 | 6/5/2019 | Environmental | VanB | 796 | A | SRR28039572 | | JBAKSY000000000 |
| 4WA10 | 6/5/2019 | Environmental | VanB | 78 | C | SRR28039571 | | JBAKSX000000000 |
| 4WA14 | 6/5/2019 | Environmental | VanB | 78 | C | SRR28039570 | | JBAKSW000000000 |
| 4WA16 | 6/5/2019 | Environmental | VanB | 796 | A | SRR28039569 | | JBAKSV000000000 |

| | | | | | | | | |
|---|---|---|---|---|---|---|---|---|
| 4WA17 | 6/5/2019 | Environmental | VanB | 78 | C | SRR28039568 | | JBAKSU000000000 |
| 4WA28B | 6/5/2019 | Environmental | VanB | 78 | C | SRR28039567 | | JBAKST000000000 |
| 4WA31 | 6/5/2019 | Environmental | VanB | 78 | C | SRR28039566 | | JBAKSS000000000 |
| 5EA26 | 29/4/2019 | Environmental | VanB | 78 | C | SRR28039565 | | JBAKSR000000000 |
| 5EA27 | 29/4/2019 | Environmental | VanA | 262 | E | SRR28039563 | | JBAKSQ000000000 |
| 5EA29 | 29/4/2019 | Environmental | VanA | 262 | E | SRR28039562 | | JBAKSP000000000 |
| 5EA30 | 29/4/2019 | Environmental | VanB | 78 | C | SRR28039561 | | JBAKSO000000000 |
| 5EA31 | 29/4/2019 | Environmental | VanA and VanB | 796 | A | SRR28039560 | | JBAKSN000000000 |
| 5EA32 | 29/4/2019 | Environmental | VanB | 78 | C | SRR28039559 | | JBAKSM000000000 |
| 5EA33 | 29/4/2019 | Environmental | VanB | 796 | A | SRR28039558 | | JBAKSL000000000 |
| 5EA34 | 29/4/2019 | Environmental | VanB | 796 | A | SRR28039557 | | JBAKSK000000000 |
| 6WE04 | 6/5/2019 | Environmental | VanA | 1421 | F | SRR28039556 | SRR28039474 | JBAKSJ000000000 |
| 6WE05 | 6/5/2019 | Environmental | VanB | 78 | C | SRR28039453 | | JBAKSI000000000 |
| 6WE11 | 6/5/2019 | Environmental | None | 80 | G | SRR28039452 | SRR28039473 | JBAKSH000000000 |
| 6WE17 | 6/5/2019 | Environmental | VanA | 1421 | F | SRR28039450 | | JBAKSG000000000 |
| 6WE18A | 6/5/2019 | Environmental | None | 80 | G | SRR28039449 | | JBAKSF000000000 |
| 6WE28 | 6/5/2019 | Environmental | VanA | 1421 | F | SRR28039448 | | JBAKSE000000000 |
| 6WE29 | 6/5/2019 | Environmental | VanB | 78 | C | SRR28039447 | | JBAKSD000000000 |
| 6WE32 | 6/5/2019 | Environmental | VanB | 796 | A | SRR28039446 | | JBAKSC000000000 |
| 6WE35 | 6/5/2019 | Environmental | VanB | 796 | A | SRR28039445 | | JBAKSB000000000 |
| 6WE36A | 6/5/2019 | Environmental | VanB | 555 | H | SRR28039444 | SRR28039472 | JBAKSA000000000 |
| 7EA02 | 6/5/2019 | Environmental | VanB | 796 | A | SRR28039443 | | JBAKRZ000000000 |
| 7EA05 | 6/5/2019 | Environmental | VanB | 796 | A | SRR28039442 | | JBAKRY000000000 |
| 7EA12 | 6/5/2019 | Environmental | VanB | 796 | A | SRR28039441 | | JBAKRX000000000 |
| 7EA19 | 6/5/2019 | Environmental | VanA | 1424 | B | SRR28039439 | | JBAKRW000000000 |
| 7EA29 | 6/5/2019 | Environmental | VanA | 80 | I | SRR28039438 | SRR28039471 | JBAKRV000000000 |
| 7EA30 | 6/5/2019 | Environmental | VanA | 80 | I | SRR28039437 | | JBAKRU000000000 |
| 7EA35 | 6/5/2019 | Environmental | VanB | 78 | C | SRR28039436 | | JBAKRT000000000 |
| 7WE01 | 29/5/2019 | Environmental | VanB | 796 | A | SRR28039435 | | JBAKRS000000000 |
| 7WE02 | 29/5/2019 | Environmental | VanB | 796 | A | SRR28039434 | | JBAKRR000000000 |

| | | | | | | | | |
|---|---|---|---|---|---|---|---|---|
| 7WE03 | 29/5/2019 | Environmental | VanB | 796 | A | SRR28039433 | | JBBDHA000000000 |
| 7WE09 | 29/5/2019 | Environmental | VanB | 796 | A | SRR28039432 | | JBAKRQ000000000 |
| 7WE11 | 29/5/2019 | Environmental | VanB | 796 | A | SRR28039431 | | JBAKRP000000000 |
| 7WE12 | 29/5/2019 | Environmental | VanB | 796 | A | SRR28039430 | | JBAKRO000000000 |
| 7WE13 | 29/5/2019 | Environmental | VanB | 796 | A | SRR28039428 | | JBAKRN000000000 |
| 7WE14 | 29/5/2019 | Environmental | VanB | 796 | A | SRR28039427 | | JBAKRM000000000 |
| 7WE15 | 29/5/2019 | Environmental | VanB | 796 | A | SRR28039426 | | JBAKRL000000000 |
| 7WE16 | 29/5/2019 | Environmental | VanB | 796 | A | SRR28039425 | | JBAKRK000000000 |
| 7WE19 | 29/5/2019 | Environmental | VanB | 796 | A | SRR28039424 | | JBAKRJ000000000 |
| AH18K015 | 7/11/2018 | Clinical | VanB | 796 | A | SRR28039423 | SRR28039469 | JBAKRI000000000 |
| AH18K037 | 11/11/2018 | Clinical | VanA | 1424 | B | SRR28039422 | SRR28039468 | JBAKRH000000000 |
| AH18K046 | 14/11/2018 | Clinical | VanA | 1421 | Q | SRR28039523 | SRR28039467 | JBAKRG000000000 |
| AH18K058 | 19/11/2018 | Clinical | VanA | 262 | E | SRR28039522 | SRR28039466 | JBAKRF000000000 |
| AH18L008 | 6/12/2018 | Clinical | VanA | 1424 | B | SRR28039521 | | JBAKRE000000000 |
| AH18L019 | 9/12/2018 | Clinical | VanB | 796 | A | SRR28039519 | | JBAKRD000000000 |
| AH18L053 | 19/12/2018 | Clinical | None | 1489 | R | SRR28039518 | SRR28039465 | JBAKRC000000000 |
| AH18L071 | 26/12/2018 | Clinical | VanB | 78 | C | SRR28039517 | SRR28039464 | JBAKRB000000000 |
| AH19A002 | 31/12/2018 | Clinical | VanB | 796 | A | SRR28039516 | | JBAKRA000000000 |
| AH19A045 | 12/1/2019 | Clinical | None | 192 | S | SRR28039515 | SRR28039463 | JBAKQZ000000000 |
| AH19A061 | 17/1/2019 | Clinical | VanB | 796 | A | SRR28039514 | | JBAKQY000000000 |
| AH19A067 | 21/1/2019 | Clinical | None | 1424 | B | SRR28039513 | | JBAKQX000000000 |
| AH19A086 | 25/1/2019 | Clinical | VanB | 796 | A | SRR28039512 | | JBAKQW000000000 |
| AH19B009 | 6/2/2019 | Clinical | VanB | 796 | A | SRR28039511 | | JBAKQV000000000 |
| AH19B023 | 10/2/2019 | Clinical | VanA | 1421 | L | SRR28039510 | SRR28039462 | JBAKQU000000000 |
| AH19B031 | 12/2/2019 | Clinical | VanA | 1421 | T | SRR28039508 | SRR28039461 | JBAKQT000000000 |
| AH19B056 | 20/2/2019 | Clinical | VanA | 262 | E | SRR28039507 | | JBAKQS000000000 |
| AH19B080 | 26/2/2019 | Clinical | VanA | 1421 | J | SRR28039506 | SRR28039460 | JBAKQR000000000 |
| AH19C003 | 1/3/2019 | Clinical | VanA and VanB | 796 | A | SRR28039505 | | JBAKQQ000000000 |
| AH19C007 | 2/3/2019 | Clinical | VanA and VanB | 796 | A | SRR28039504 | | JBAKQP000000000 |
| AH19C045 | 11/3/2019 | Clinical | VanA | 1424 | B | SRR28039503 | | JBAKQO000000000 |

| | | | | | | | | |
|---|---|---|---|---|---|---|---|---|
| AH19C052 | 14/3/2019 | Clinical | VanA | 1421 | J | SRR28039502 | | JBAKQN000000000 |
| AH19C054 | 14/3/2019 | Clinical | VanA | 1424 | B | SRR28039501 | | JBAKQM000000000 |
| AH19C059 | 16/3/2019 | Clinical | VanA | 1424 | B | SRR28039500 | | JBAKQL000000000 |
| AH19C096 | 29/3/2019 | Clinical | None | 252 | U | SRR28039499 | SRR28039458 | JBAKQK000000000 |
| AH19C103 | 31/3/2019 | Clinical | VanB | 78 | C | SRR28039497 | | JBAKQJ000000000 |
| AH19D045 | 19/4/2019 | Clinical | None | 80 | V | SRR28039496 | SRR28039588 | JBAKQI000000000 |
| AH19D049 | 20/4/2019 | Clinical | VanB | 252 | K | SRR28039495 | SRR28039587 | JBAKQH000000000 |
| AH19D061 | 22/4/2019 | Clinical | VanB | 796 | A | SRR28039494 | | JBAKQG000000000 |
| AH19E019 | 9/5/2019 | Clinical | VanB | 796 | A | SRR28039493 | | JBAKQF000000000 |
| AH19E044 | 21/5/2019 | Clinical | VanA | 1421 | L | SRR28039492 | | JBAKQE000000000 |
| AH19E068 | 27/5/2019 | Clinical | VanB | 78 | C | SRR28039457 | | JBAKQD000000000 |
| AH19F003 | 1/6/2019 | Clinical | None | 1424 | W | SRR28039456 | SRR28039586 | JBAKQC000000000 |
| AH19F005 | 2/6/2019 | Clinical | VanB | 796 | A | SRR28039455 | | JBAKQB000000000 |
| AH19F029 | 7/6/2019 | Clinical | VanB | 80 | M | SRR28039454 | SRR28039585 | JBAKQA000000000 |
| AH19F040 | 12/6/2019 | Clinical | VanB | 796 | A | SRR28039589 | | JBAKPZ000000000 |
| AH19F067 | 25/6/2019 | Clinical | VanB | 796 | N | SRR28039555 | SRR28039584 | JBAKPY000000000 |
| AH19F070 | 26/6/2019 | Clinical | VanB | 796 | N | SRR28039554 | | JBAKPX000000000 |
| AH19F071 | 26/6/2019 | Clinical | VanB | 796 | A | SRR28039553 | | JBAKPW000000000 |
| AH19F078 | 27/6/2019 | Clinical | VanB | 796 | A | SRR28039552 | | JBAKPV000000000 |
| AH19G039 | 13/7/2019 | Clinical | VanB | 796 | A | SRR28039551 | | JBAKPU000000000 |
| AH19G046 | 16/7/2019 | Clinical | VanB | 796 | A | SRR28039550 | | JBAKPT000000000 |
| AH19G054 | 21/7/2019 | Clinical | VanA | 1425 | O | SRR28039549 | SRR28039583 | JBAKPS000000000 |
| AH19G064 | 25/7/2019 | Clinical | VanA | 1424 | B | SRR28039548 | | JBAKPR000000000 |
| AH19G077 | 29/7/2019 | Clinical | VanA | 1424 | B | SRR28039547 | | JBAKPQ000000000 |
| AH19G079 | 30/7/2019 | Clinical | VanA | 1424 | B | SRR28039545 | | JBAKPP000000000 |
| AH19H004 | 2/8/2019 | Clinical | VanA | 1424 | B | SRR28039544 | | JBAKPO000000000 |
| AH19H005 | 3/8/2019 | Clinical | VanA | 1424 | B | SRR28039543 | | JBAKPN000000000 |
| AH19H014 | 6/8/2019 | Clinical | VanA | 1424 | B | SRR28039542 | | JBAKPM000000000 |
| AH19H029 | 10/8/2019 | Clinical | VanB | 796 | A | SRR28039541 | | JBAKPL000000000 |
| AH19H030 | 11/8/2019 | Clinical | VanA | 1424 | B | SRR28039540 | | JBAKPK000000000 |

| | | | | | | | | |
|---|---|---|---|---|---|---|---|---|
| AH19H058 | 21/8/2019 | Clinical | None | 1283 | X | SRR28039539 | SRR28039582 | JBAKPJ000000000 |
| AH19H062 | 22/8/2019 | Clinical | VanA | 1424 | B | SRR28039538 | | JBAKPI000000000 |
| AH19H067 | 24/8/2019 | Clinical | None | 212 | Y | SRR28039537 | SRR28039581 | JBAKPH000000000 |
| AH19H074 | 27/8/2019 | Clinical | VanA | 1424 | B | SRR28039536 | | JBAKPG000000000 |
| AH19H083 | 29/8/2019 | Clinical | VanB | 796 | A | SRR28039534 | | JBAKPF000000000 |
| AH19I021 | 10/9/2019 | Clinical | VanA | 1424 | B | SRR28039533 | | JBAKPE000000000 |
| AH19I027 | 11/9/2019 | Clinical | VanB | 796 | A | SRR28039532 | | JBAKPD000000000 |
| AH19I041 | 18/9/2019 | Clinical | VanA | 1424 | B | SRR28039531 | | JBAKPC000000000 |
| AH19I049 | 20/9/2019 | Clinical | VanA | 1424 | B | SRR28039530 | | JBAKPB000000000 |
| AH19I064 | 25/9/2019 | Clinical | VanB | 796 | A | SRR28039529 | | JBAKPA000000000 |
| AH19J002 | 2/10/2019 | Clinical | VanB | 796 | A | SRR28039528 | | JBAKOZ000000000 |
| AH19J027 | 17/10/2019 | Clinical | VanB | 78 | C | SRR28039527 | | JBAKOY000000000 |
| AH19J038 | 20/10/2019 | Clinical | VanA | 1424 | B | SRR28039526 | | JBAKOX000000000 |
| AH19K007 | 2/11/2019 | Clinical | None | 1425 | O | SRR28039525 | | JBAKOW000000000 |
| AH19K008 | 4/11/2019 | Clinical | None | 1424 | B | SRR28039491 | | JBAKOV000000000 |
| AH19K011 | 4/11/2019 | Clinical | VanA | 1424 | B | SRR28039490 | | JBAKOU000000000 |
| AH19K039 | 12/11/2019 | Clinical | VanB | 796 | A | SRR28039489 | | JBAKOT000000000 |
| AH19K042 | 14/11/2019 | Clinical | VanB | 78 | C | SRR28039488 | | JBAKOS000000000 |
| AH19K055 | 18/11/2019 | Clinical | VanA | 1424 | B | SRR28039487 | | JBAKOR000000000 |
| AH19K073 | 25/11/2019 | Clinical | VanA | 1424 | B | SRR28039486 | | JBAKOQ000000000 |
| AH19L034 | 13/12/2019 | Clinical | None | 1913 | Z | SRR28039485 | SRR28039580 | JBAKOP000000000 |
| Bed01 | 6/5/2019 | Environmental | None | 696 | P | SRR28039484 | SRR28039578 | JBAKOO000000000 |
| Bed08 | 6/5/2019 | Environmental | VanB | 78 | C | SRR28039483 | | JBAKON000000000 |
| FFH15 | 21/5/2019 | Environmental | VanB | 796 | A | SRR28039482 | | JBBDGZ000000000 |
| ICU20 | 13/5/2019 | Environmental | VanB | 796 | A | SRR28039480 | | JBAKOM000000000 |
| ICU55 | 13/5/2019 | Environmental | VanB | 796 | A | SRR28039479 | | JBAKOL000000000 |
| ICU56 | 13/5/2019 | Environmental | VanB | 796 | A | SRR28039478 | | JBAKOK000000000 |
| ICU61A | 13/5/2019 | Environmental | VanB | 796 | A | SRR28039477 | | JBAKOJ000000000 |
| ICU64 | 13/5/2019 | Environmental | VanB | 796 | A | SRR28039476 | | JBAKOI000000000 |

**Supp. Table 2 – Summary of putative *Enterococcus faecium* genomic transmission clusters**

| Putative genomic transmission cluster | Total genomes | Clinical | Environmental | *vanA* | *vanB* | *vanA* and *vanB* | No *van* operon |
|---|---|---|---|---|---|---|---|
| 1 | 32 | 9 | 23 | 0 | 31 | 1 | 0 |
| 2 | 26 | 22 | 4 | 25 | 0 | 0 | 1 |
| 3 | 14 | 2 | 12 | 0 | 14 | 0 | 0 |
| 4 | 10 | 3 | 7 | 0 | 10 | 0 | 0 |
| 5 | 6 | 2 | 4 | 0 | 6 | 0 | 0 |
| 6 | 4 | 0 | 4 | 0 | 4 | 0 | 0 |
| 7 | 3 | 0 | 3 | 3 | 0 | 0 | 0 |
| 8 | 2 | 1 | 1 | 0 | 2 | 0 | 0 |
| 9 | 2 | 1 | 1 | 0 | 2 | 0 | 0 |
| 10 | 2 | 2 | 0 | 0 | 0 | 2 | 0 |
| 11 | 2 | 2 | 0 | 2 | 0 | 0 | 0 |
| 12 | 2 | 2 | 0 | 0 | 2 | 0 | 0 |
| 13 | 2 | 2 | 0 | 1 | 0 | 0 | 1 |
| 14 | 2 | 2 | 0 | 2 | 0 | 0 | 0 |
